# Mapping the Association between Mental Health and People’s Perceived and Actual Ability to Practice Hygiene-Related Behaviours in Humanitarian and Pandemic Crises: A Scoping Review

**DOI:** 10.1101/2023.05.18.23290179

**Authors:** Emily Yasmin Ghassemi, Astrid Hasund Thorseth, Karine Le Roch, Thomas Heath, Sian White

## Abstract

Humanitarian crises such as disease outbreaks, conflict and displacement and natural disasters affect millions of people primarily in low- and middle-income countries. Here, they often reside in areas with poor environmental health conditions leading to an increased burden of infectious diseases such as diarrheal and respiratory infections. Water, sanitation, and hygiene behaviours are critical to prevent such infections and deaths.

A scoping review was conducted to map out what is known about the association between three mental health issues and people’s perceived and actual ability to practice hygiene-related behaviours, particularly handwashing, in humanitarian and pandemic crises. Published and grey literature was identified through database searches, humanitarian-relevant portals, and consultations with key stakeholders in the humanitarian sector.

25 publications were included, 21 were peer-reviewed published articles and four were grey literature publications. Most of the studies were conducted in China (n=12) and most were conducted in a pandemic outbreak setting (n=20). Six studies found a positive correlation between handwashing and anxiety where participants with higher rates of anxiety were more likely to practice handwashing with soap. Four studies found an inverse relationship where those with higher rates of anxiety were less likely to wash their hands with soap. The review found mixed results for the association between handwashing and depression, with four of the seven studies reporting those with higher rates of depression were less likely to wash their hands, while the remaining studies found that higher depressions scores resulted in more handwashing. Mixed results were also found between post-traumatic stress disorder (PTSD) and handwashing. Two studies found that lower scores of PTSD were associated with better hygiene practices, including handwashing with soap.

The contradictory patterns suggest that researchers and practitioners need to explore this association further, in a wider range of crises, and need to standardize tools to do so.

## Introduction

Humanitarian crises pose a serious threat to the health, safety, security, and wellbeing of communities, and affect large numbers of people across the world [1]. Crises can lead to forced displacement and disruptions to physical and social environments. Therefore, for security reasons, crisis-affected populations often live in densely populated environments with inadequate sanitation and hygiene leading to an increased risk of outbreak-related diseases, and diarrhoeal and respiratory illnesses [2-6]. Consequently, a core component of humanitarian responses to crises often involves water, sanitation, and hygiene (WASH) interventions [3, 7]. In particular, improving hand hygiene behaviour, is known to be highly cost-effective and can result in diarrhoeal disease reductions of 23% - 48% and reductions in respiratory infections of 21-23% [8-13].

However, handwashing behaviour is embedded in daily routines and is an activity that demands effort, time, and motivation. Like most behaviours, handwashing is known to be influenced by a range of cognitive determinants and factors in the social and physical environment [14]. Handwashing may therefore be more challenging for crisis-affected populations to practice given that they are more likely to have been exposed to traumatic situations and face difficult living conditions, all of which may affect their mental health.

Mental health is more than just the absence of a mental disorder [15]. It is a state of well-being, defined as an individual being able to realise their potential, cope with the everyday stresses of life, work productively, and contribute to their community. In humanitarian crises, depressive and anxiety disorders are the most common mental health issues encountered [16, 17]. Additionally, studies have shown that traumatic disorders, particularly post-traumatic stress disorders (PTSD) and obsessive-compulsive disorders (OCD) related to trauma, also occur more frequently during humanitarian crises [16-19].

The link between handwashing practices and mental health has yet to be extensively studied [20]. However, literature from high-income settings indicates that some personal hygiene behaviours, including handwashing, may be affected by mental health. For example, poor personal hygiene is known to be a feature of certain psychiatric conditions such as schizophrenia and depression [21-23]. The lack of research on this topic in low- or middle-income countries (LMICs) or humanitarian crises may be partially because, in such settings, mental health is often under resourced and staffed, and culturally stigmatised, resulting in mental health issues going undiagnosed and untreated [20]. Similarly, there are often barriers to understanding handwashing behaviour in these settings. Self-reported handwashing behaviour is likely to overestimate actual practice because handwashing is a socially desirable behaviour [24, 25]. Observation, which is considered a more reliable method of measuring handwashing, is time consuming and resource intensive and is therefore rarely undertaken at scale [14, 26].

This scoping review aims to map out what is known about the association between mental health and the perceived and actual ability of adults to practice handwashing in humanitarian crises. It does so with the aim of identifying opportunities for humanitarian practitioners or researchers to better integrate mental health and hygiene work to improve the wellbeing and health of crisis-affected populations.

## Methods

A protocol for the scoping review was drafted following the process outlined by Arksey and O’Malley [27]. This process involves five steps: 1) Identifying relevant studies through a clear search strategy, 2) a ‘Consultation exercise’ to seek out grey literature for inclusion, 3) Study selection based on eligibility criteria, 4) Charting the data, 5) Collating, summarizing, and reporting the results. These steps are described below. The scoping review protocol is available on request from the corresponding author.

### Key definitions, search terms and search strategy for published literature

To undertake the review three main parameters, and associated sub-concepts within these, had to be defined and search terms identified. These main parameters included humanitarian crises affecting LMICs, mental health, and handwashing. To arrive at these definitions and terms an exploratory search of the literature was conducted and then this was refined in discussion with stakeholders who worked on hygiene or mental health programmes in humanitarian settings. The parameters, definitions, terms and a rationale for their selection are provided in Table 1 and in detail in S1 Table.

**Table 1:**
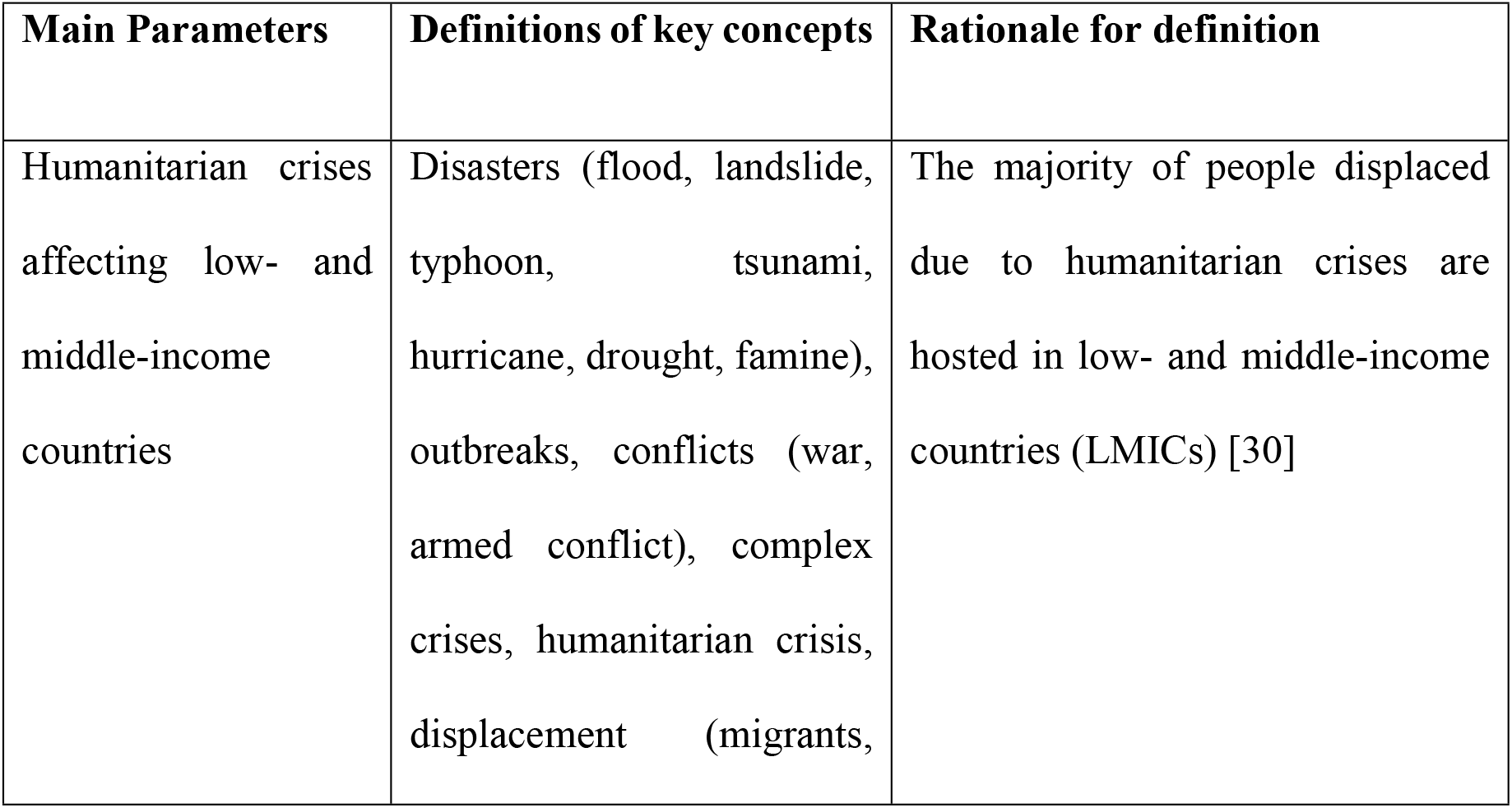
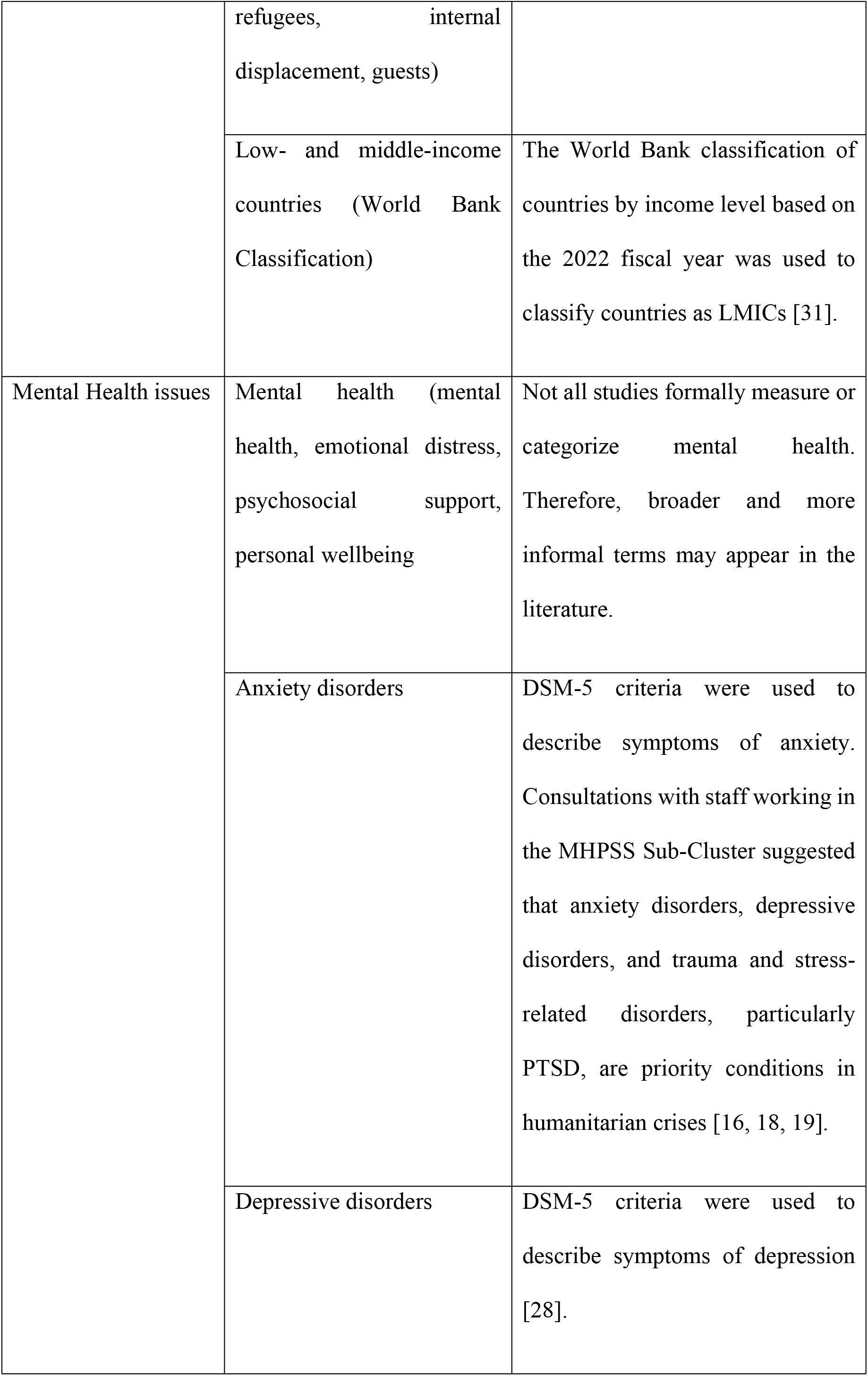
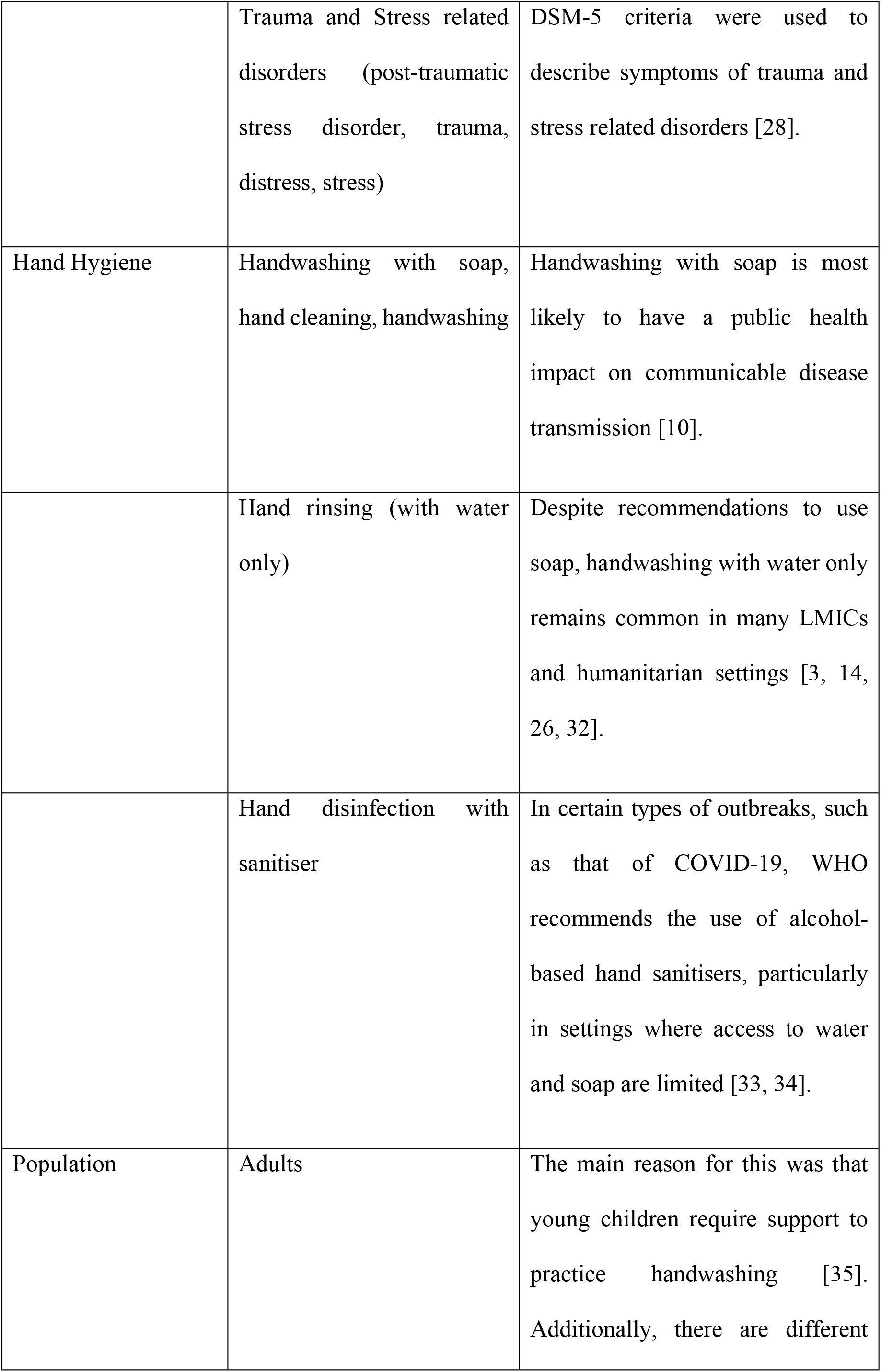
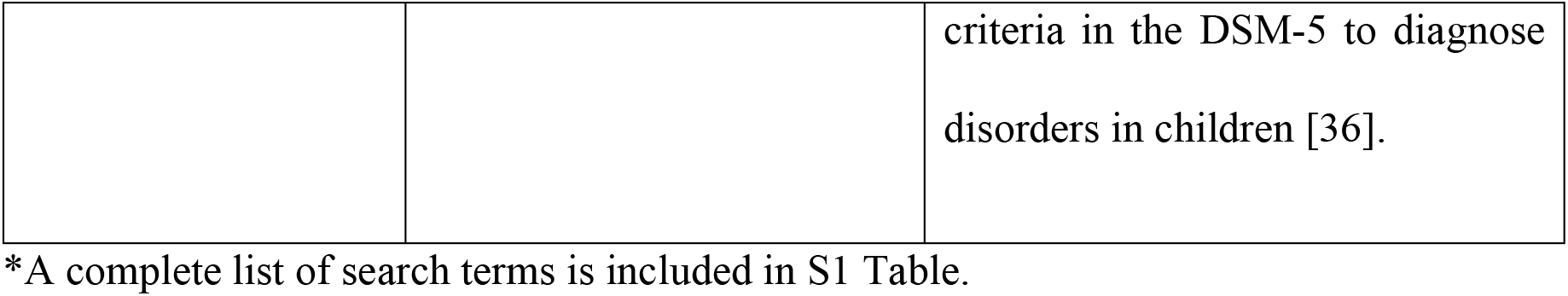
A summary of the main parameters, their definitions and search terms and a rationale for their inclusion

| Main Parameters | Definitions of key concepts | Rationale for definition |
| --- | --- | --- |
| Humanitarian crises affecting low- and middle-income countries | Disasters (flood, landslide, typhoon, tsunami, hurricane, drought, famine), outbreaks, conflicts (war, armed conflict), complex crises, humanitarian crisis, displacement (migrants, | The majority of people displaced due to humanitarian crises are hosted in low- and middle-income countries (LMICs) [30] |
|  | refugees, internal displacement, guests) |  |
|  | Low- and middle-income countries (World Bank Classification) | The World Bank classification of countries by income level based on the 2022 fiscal year was used to classify countries as LMICs [31]. |
| Mental Health issues | Mental health (mental health, emotional distress, psychosocial support, personal wellbeing | Not all studies formally measure or categorize mental health. Therefore, broader and more informal terms may appear in the literature. |
|  | Anxiety disorders | DSM-5 criteria were used to describe symptoms of anxiety. Consultations with staff working in the MHPSS Sub-Cluster suggested that anxiety disorders, depressive disorders, and trauma and stress-related disorders, particularly PTSD, are priority conditions in humanitarian crises [16, 18, 19]. |
|  | Depressive disorders | DSM-5 criteria were used to describe symptoms of depression [28]. |
|  | Trauma and Stress related disorders (post-traumatic stress disorder, trauma, distress, stress) | DSM-5 criteria were used to describe symptoms of trauma and stress related disorders [28]. |
| Hand Hygiene | Handwashing with soap, hand cleaning, handwashing | Handwashing with soap is most likely to have a public health impact on communicable disease transmission [10]. |
|  | Hand rinsing (with water only) | Despite recommendations to use soap, handwashing with water only remains common in many LMICs and humanitarian settings [3, 14, 26, 32]. |
|  | Hand disinfection with sanitiser | In certain types of outbreaks, such as that of COVID-19, WHO recommends the use of alcohol-based hand sanitisers, particularly in settings where access to water and soap are limited [33, 34]. |
| Population | Adults | The main reason for this was that young children require support to practice handwashing [35]. Additionally, there are different |
|  |  | criteria in the DSM-5 to diagnose disorders in children [36]. |

\*A complete list of search terms is included in S1 Table.

**Table 2.** Type of humanitarian crisis and the study setting.

| Type of humanitarian crisis and setting | Count (n = 25) |
| --- | --- |
| Type of crisis |  |
| Outbreak | 20 |
| Complex Crisis | 2 |
| Conflict | 1 |
| Humanitarian emergency (general) | 2 |
| Natural disaster | 0 |
| Country |  |
| China | 12 |
| Ethiopia | 3 |
| Multiple countries | 3 |
| Malawi | 2 |
| Iraq | 2 |
| Iran | 1 |
| Sierra Leone | 1 |
| Peru | 1 |
| Type of Setting |  |
| Urban | 14 |
| Rural | 3 |
| Both | 4 |
| Not specified | 4 |
| Studied Population |  |
| Adults (general) | 17 |
| Displaced populations (refugees or IDPs) | 4 |
| Factory workers | 2 |
| University students | 1 |
| Pregnant women | 1 |

LMICs were defined according to the World Bank Classifications and humanitarian crises was defined as covering disasters, outbreaks, conflicts, and complex crises. Mental health was defined through consultations with staff working within the Mental Health and Psychosocial Support (MHPSS) Sub-Cluster who suggested that priority mental health conditions in humanitarian settings are trauma and stress-related disorders, particularly PTSD; depressive disorders, and anxiety disorders [16, 18, 19]. Accordingly, these were the primary mental health issues researched with each concept being defined according to Diagnostic and Statistical Manual of Mental Health Disorders (DSM-5) criteria [28]. Discussions with people in the MHPSS sector revealed that OCD was rarely assessed in humanitarian contexts, and it was therefore not included. Hand hygiene included concepts related to ‘hygiene behaviour’. Most literature focuses on handwashing, specifically. Handwashing with soap, hand rinsing with water only and hand sanitising is most likely to have a public health impact on communicable disease transmission in crises and is commonly the focus of hygiene promotion programs in these settings [10]. For that reason, the review focused narrowly on handwashing behaviour. Handwashing was defined as handwashing with soap, hand cleaning (with water), or hand disinfection with sanitiser. Handwashing with ash was excluded given that evidence of its effectiveness at removing or killing pathogens is limited [29]. For each parameter and sub-concept Medical Subject Headings (MeSH) terms were used along with other targeted keywords and search terms.

Five electronic databases were initially searched in June and July 2021: PubMed, Medline, Global Health, Embase, and the online library of the London School of Hygiene and Tropical Medicine (LSHTM). The search was repeated in January 2023 and covered July 2021 until January 2023, capturing literature published after the initial search.

### Identification of grey literature

Grey literature was also identified in June and July 2020 through a two-step process. This initially involved searching publicly available humanitarian or hygiene-related websites, including ReliefWeb, and resources on the websites of the Global WASH Cluster, The Global handwashing Partnership, the MHPSS Sub-Cluster, the United Nations Children’s Fund (UNICEF), the United Nations High Commissioner for Refugees (UNHCR), the Inter-Agency Standing Committee (IASC), the International Committee of the Red Cross (ICRC), the International Federation of the Red Cros (IFRC), the International Rescue Committee (IRC), Action contre la Faim (ACF), and the Centers for Disease Control and Prevention (CDC). The WASH and mental health resource pages were scanned in particular. The grey literature searches used a combination of the terms and MeSH terms mental health, hand hygiene, and humanitarian crises, to identify resources (Table 1).

The second process employed to identify grey literature was a consultation process [27]. This involved contacting academics who have done research on relevant topics and humanitarian practitioners and contacting organizations within the Global Handwashing Partnership, the Hand Hygiene for all Initiative, the Global WASH Cluster, the MHPSS Sub-Cluster, and the IASC Reference Group and asking them to share any relevant grey literature documents that they were aware of. Requests for literature were also made via several public forums including the Sustainable Sanitation Alliance (SuSanA) and several humanitarian and WASH oriented Facebook groups.

### Inclusion and exclusion criteria

No limit was placed on the publication date of published or grey literature and all study designs were included. Therefore, any original research studies, policy briefs, theoretical papers, and qualitative or quantitative data were included. To be eligible, publications had to mention handwashing and mental health in the results of the manuscript or findings sections of grey literature documents and publication had to be in English.

### Data Extraction and Analysis

Publications identified through the search process were imported into EndNote 20 and duplications were removed. Publications were initially screened based on their titles and abstracts, and subsequently full texts reads were done to assess whether they met the eligibility criteria. The charting process for included publications involved summarising the following information for each document: journal or publishing organization, year of publication, study location, aims and objectives, description of context, study population, description of methods, outcome measures (for both the mental health and handwashing), description of association between mental health and handwashing, and a summary of any identified opportunities for sector integration or programme strengthening. Patterns across the study parameters were then identified and descriptively summarised.

### Ethics statement

This study was approved by the Research Ethics Committee at the London School of Hygiene and Tropical Medicine (submission ID: 25636). No consent was required given that this study did not involve human subjects.

## Results

The search of published literature yielded 3,234 texts. After removing duplicates and conducting a screening of titles, 784 studies remained. A more detailed screening of abstracts led to 186 full texts being reviewed. Of these, 21 met the inclusion criteria. The grey literature search and the consultations with relevant experts led to the identification of 54 documents, of which four were eligible for inclusion. A PRISMA flow chart summarising the data screening process is provided in Fig 1.

**Fig 1.**
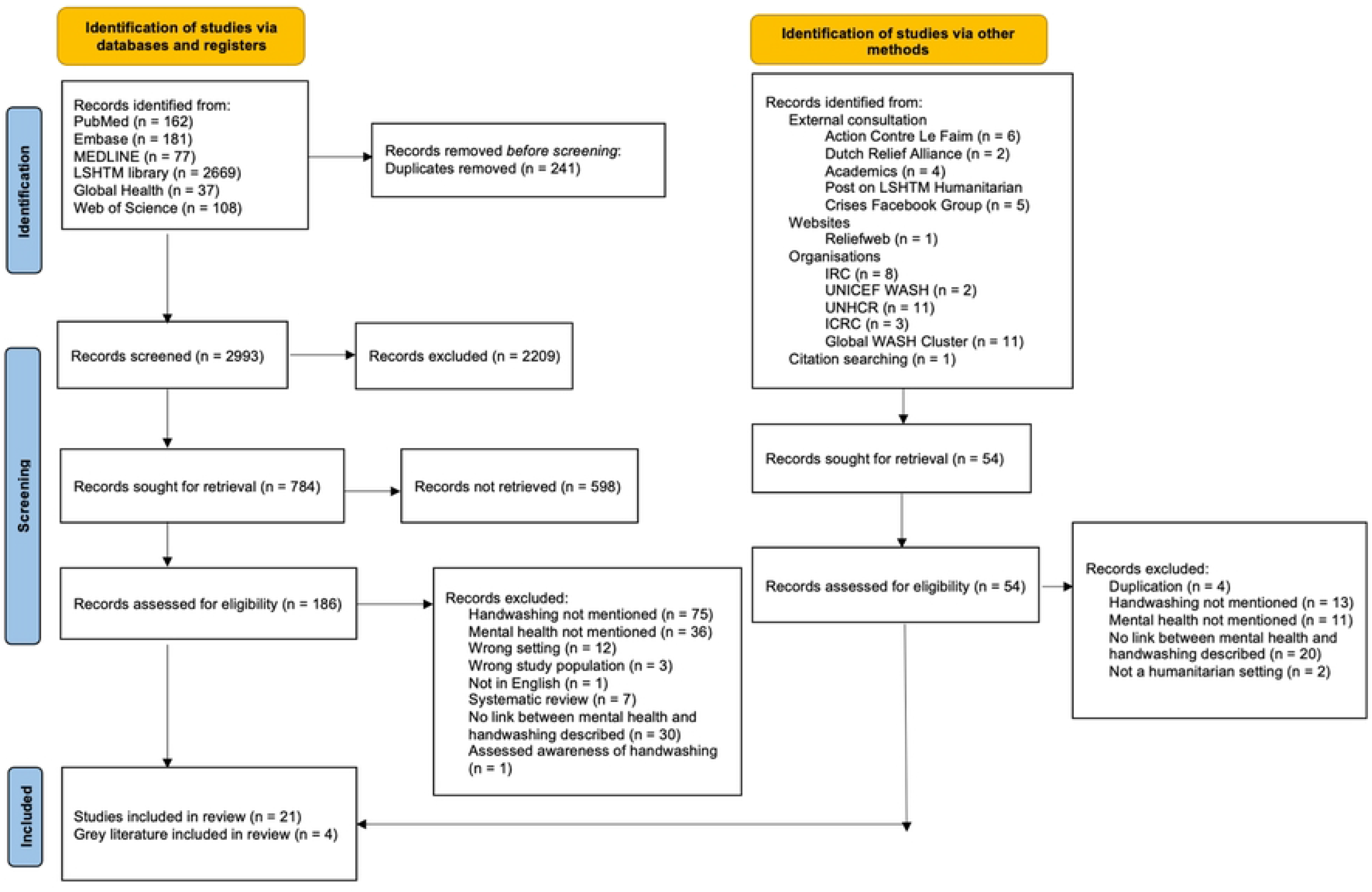
PRISMA flow diagram. This diagram outlines the screening process.

### Characteristics of the included studies

All the included manuscripts or documents were published after 2013, with one published in 2013, one in 2014, one in 2016, 11 in 2020, eight in 2021, and three in 2022. The majority of the publications in the last three years were focused on or mentioned the COVID-19 pandemic.

Table 4 provides a summary of the study settings and study populations for the included publications. Most publications (n = 20) reported on disease outbreaks, particularly COVID-19, Ebola Virus Disease (EVD) (n = 1), and Avian Influenza A (H7N9) (n = 1). COVID-19 studies were mostly conducted in China (n = 12) and Ethiopia (n = 3). Most grey literature documents (n = 3) provided data on multiple countries. The studies conducted in Malawi, Sierra Leone, and Iraq were conducted during a complex crisis, EVD outbreak, and conflict, respectively. No included publications were set in a natural disaster setting. Of the country-specific studies, most were conducted in urban China (n = 9) and one in an urban region of Iran (n = 1). Only three studies were conducted in rural areas. Four studies investigated urban and rural settings in China (n = 3) and Sierra Leone (n = 1). Three studies did not specify a setting type. Most publications (n = 17) studied adults in general, but some focused more narrowly on sub-sections of the population such as factory workers (n = 2), displaced populations (n = 4) or pregnant women (n = 1). The grey literature documents (n = 3) did not explicitly describe study populations.

The majority of included peer-reviewed publications featured a cross-sectional study design (n = 19). Two papers described a longitudinal design that incorporated data collection from the same households before and after handwashing intervention. 15 of the studies used questionnaires as their primary data collection method, one used interviews, four studies conducted both surveys and interviews, and one study used observations, focus group discussions, in-depth interviews and handwashing demonstrations. The grey literature were all guidance documents featuring case studies.

### Outcome measures

Eight studies focused on a particular mental health condition. Anxiety was the primary outcome in five publications [37-41]. The Generalized Anxiety Disorder 7-item scale (GAD-7) was used to measure anxiety in two studies [37, 39]. One publication used the Self-rating Anxiety Scale (SAS) to measure anxiety [40] and another study used the COVID-19 Induced Anxiety Scale (CIAS) for the same purpose [41]. Other study-specific questions addressing worry, life and death pondering, difficulty in daily life, and anxiety related to perceived infection probability were used to measure anxiety in one publication [38].

Depression was measured in three studies. The Patient Health Questionnaire-9 (PHQ-9) was used to assess this in all three studies [42-44].

No included publications had PTSD as the sole mental health outcome.

13 studies assessed more than one mental health outcome. Anxiety, depression, and PTSD were the outcomes of seven studies [45-47, 49-52]. Anxiety, depression, and PTSD were jointly assessed through the Hopkins Symptom Checklist (HSCL-25) in one study [47]. Three other studies used the Depression, Anxiety, and Stress scale-21 (DASS) to collect data on these disorders [48-50]. Additionally, one study referred to anxiety, depression, and trauma as “common mental disorders” [46[p350]] (CMDs) experienced by vulnerable populations in developing countries exposed to emergencies. CMDs were measured through the WHO’s Self-Reporting Questionnaire (SRQ-20). Another study also relied on the SRQ-20 but referred to the mental health conditions of anxiety, depression, and stress. Lastly, in two studies mental health was described as psychological distress [53] or psychological well-being [51] measured by the Kessler-10 questionnaire and the General Health Questionnaire (GHQ-12), respectively. Three grey literature documents focused on a range of mental health issues that commonly arise in crises or outbreaks and did not describe specific tools for measuring these but suggested that mental health conditions should be assessed by trained mental health experts [45, 54, 55]. Anxiety and depression were the outcomes of four studies [56-59]. The GAD-7 was used to measure anxiety in three studies [56, 57, 59], and the State-trait Anxiety Inventory was used in another [58]. To measure depression, two studies used the PHQ-9 [56, 59], one study used the PHQ-2, and one study used the Self-rating Depression Scale (SDS) [58].

Lastly, two studies were included despite not having any mental health parameters as an outcome. Both studies [60, 61], mental health arose from interviews as a challenge to practicing handwashing behaviour in displacement camps in Iraq, therefore meeting inclusion criteria.

Handwashing with soap was the most common outcome measure (n = 11). In seven studies, handwashing with soap was self-reported through questions developed specifically for that study [37, 38, 47-49, 56, 57]. One study referred to handwashing at the five key moments [52]. Self-reported questions included the average number of times people washed their hands daily with soap and running water [57], reported changes in handwashing habits [38], frequency of immediate handwashing when returning home [37], and average handwashing duration [37], among others. Three studies used self-reported questions on handwashing used in prior research, including a standardised Barrier Analysis Survey [60] and standardised surveys or questions related to hand hygiene during the COVID-19 pandemic [39, 48, 49]. Three publications used proxy measures to assess handwashing behaviour, such as the standard global handwashing indicator, which assesses the availability of handwashing facilities with soap and water present [45, 46, 60].

Three studies measured both handwashing with soap and hand sanitising as hygiene outcomes [42-44]. All three publications measured this behaviour through study-specific self-reported questionnaires with questions including the frequency at which participants sanitised their hands with soaps or alcohol-based sanitisers [43, 44] and perception of cleanliness after disinfecting and handwashing [42]. One study referred to ‘hand hygiene’ as the primary outcome [58]. While this was not clearly defined it was assumed that this may also include both handwashing with soap and hand sanitising behaviours. This behaviour was measured through a study-specific questionnaire with questions including preventive measures followed against COVID-19 infection [58]. Lastly, two grey literature publications did not specify how hygiene behaviour was defined, nor did they mention how this behaviour was measured [54, 55].

### Anxiety and handwashing

Of the 11 studies that provided data on anxiety and handwashing with soap, six found a positive correlation between handwashing with soap and anxiety, where participants with higher rates of anxiety were more likely to practice handwashing with soap [37-39, 41 56, 57]. Four studies found an inverse relationship in which those with higher rates of anxiety were less likely to wash their hands with soap [48-50, 59]. One study found no direct relationship between anxiety and handwashing [40]. No included publications reported on anxiety and hand sanitising. However, one publication reported on anxiety and hand hygiene in general [58]. This study found that respondents’ anxiety did not relate to public behaviour change and preventive measures.

During the COVID-19 pandemic, Birhanu et al. [41] found a strong positive correlation between anxiety and protective behaviours, of which frequent handwashing was one. During the outbreak in a urban setting in Ethiopia, participants who displayed high protective behaviour had twice the odds of COVID-19 induced anxiety compared to participants with a moderate level of preventive behaviour (AOR = 2.2, 95% CI: 1.5 – 3.3) [41]. Qian et al. [37] also found a positive correlation between anxiety and handwashing among adults. A significantly higher proportion of residents near the outbreak’s epicentre in Wuhan, China reported moderate or severe anxiety than participants further away from the outbreak in Shanghai, 32.8% and 20.5%, respectively. Wuhan residents were also more likely to follow the recommendations than the Shanghai residents. However, both groups reported always washing hands immediately when returning home, a new recommended critical moment for handwashing during the pandemic [37]. Similarly, in Iran, Mohammadpour et al. [39] found a borderline significant relationship (P-value = 0.06) where among participants with anxiety, for those who believed that one must practice handwashing with soap as much as possible during the COVID-19 epidemic, the mean score for anxiety was lower than the mean score for those who did not feel frequent handwashing was important. Ni and colleagues [57] also found a positive correlation between anxiety and handwashing. Here, however, a higher frequency of handwashing (Adjusted OR (aOR)=1.02, p=0.03), living near the epicenter (aOR=2.85, P-value < 0.01), and meeting the screening criteria for depression (aOR=24.20, P-value < 0.01) were independently associated with moderate and severe anxiety symptoms. Among pregnant women in China also during the COVID-19 pandemic, decreased rates of anxiety led to decreased handwashing rates (r = -0.08, P-value < 0.001) [56]. After adjusting for socio-demographic and pregnancy-related factors, only social support was associated with lower anxiety levels (aOR = 0.86-0.87) and a higher frequency of handwashing (aOR = 1.06) [56]. Similar findings were also reported in other outbreaks. In the EVD outbreak in Sierra Leone, individuals with higher reported anxiety levels reported increased EVD prevention behaviours [47]. In response to the H7N9 outbreak in China, 42.80% of respondents increased handwashing habits [38].

Among a similar study population of urban residents in China during the COVID-19 pandemic, handwashing with soap was significantly associated with lower anxiety scores [48]. During the epidemic’s peak, particularly handwashing with soap was significantly associated with lower anxiety scores among survey participants [49]. Access to adequate availability of handwashing materials in cafeterias, libraries, and classes was also linked to university students in Ethiopia being 42% less likely to develop anxiety compared to students with inadequate access to these materials (AOR = 0.58, CI: 0.43 – 0.81) [51]. Similar results were also found among study populations in rural residents in China, where high levels of anxiety predicted lower compliance levels with preventive behaviours (OR = 1.55, 95% CI: 1.10–2.18, P-value < 0.05), including frequent handwashing [59].

In a sample of 28 provinces and cities in China, hand hygiene was one of the most common preventive measures to be adhered to, with 82.4% of respondents demonstrating this. However, respondents’ anxiety did not relate to public behaviour change and preventive measures [59]. In a similar study population in China, it was found that there was no significant direct relationship between anxiety and compliance with preventive behaviours, including handwashing [40].

### Depression and handwashing

Eight studies provided data on depression and handwashing outcomes. Two studies found positive relationships where the variables of handwashing and depression increased and decreased together [56, 60]. Four studies found inverse relationships where those with higher rates of depression were more likely to wash their hands or vice versa [38, 43, 44, 60]. Following the EVD outbreak in Sierra Leone, one study found no significant association between depression and handwashing [47]. Similar results were found during the COVID-19 outbreak in China, where no statistically significant relationship was found between depression and compliance to preventive behaviours, including handwashing [59].

One publication reported on depression and hand hygiene across 28 provinces and cities in China, 82.4% of participants adhered to frequent hand hygiene practices during the COVID-19 outbreak. However, fewer respondents with depression practiced handwashing with soap than those without depression [58].

During the COVID-19 outbreak, amongst pregnant women in China, those with lower rates of depression were less likely to wash their hands with soap daily [38]. Similar results were found where participants who never felt clean after repeatedly disinfecting and scrubbing their hands and items had significantly higher depression scores (P-value < 0.001) [42]. Being calm (OR 0.344, 95% CI 0.186-0.635) and having social support (OR 0.529, 95% CI 0.308-0.908) was negatively associated with depression.

Contrastingly, among factory workers in Shenzhen, China, more exposure to unofficial web-based media was associated with higher compliance with hand sanitising and higher depressive symptoms after adjusting for demographic variables. However, exposure to information about positive outcomes for COVID-19 patients, prevention and treatment developments, and heroic stories about frontline healthcare workers were associated with better mental health and higher compliance to preventive measures, including hand sanitising (adjusted B= –0.16, P-value = 0.045) [43]. In another study in the sample population, depressive symptoms were associated with less self-reported sanitising of hands (AORs of 0.93, CI 0.91-0.94) [44]. During the epidemic’s peak, observing better hygiene practices, particularly washing hands with soap and water, and doing so after coughing, rubbing the nose, and sneezing, were significantly associated with lower depression scores [48, 49].

### PTSD and handwashing

Different relationships were found among the included publications with PTSD and handwashing outcomes. Two studies conducted during the COVID-19 pandemic found an inverse relationship. Observing better hygiene practices was significantly associated with lower scores in PTSD symptoms [48, 49]. Washing hands immediately after coughing, sneezing, or rubbing the nose was significantly associated with lower PTSD symptoms (B = - 0.47) [48]. Similarly, after the initial peak of COVID-19 cases in China, significant positive associations were found between washing hands with soap and water most of the time and occasionally and PTSD symptoms [49]. When participants washed their hands immediately after coughing, rubbing the nose, or sneezing, this was associated with lower scores of PTSD [49].

PTSD and handwashing during the COVID-19 pandemic yielded similar results to PTSD and handwashing during the EVD outbreak. A positive relationship was found in the latter, where higher war exposure levels (B = 0.45; P-value = 0.003) were associated with greater EVD prevention behaviours such as frequent handwashing. PTSD symptoms, however, were associated with fewer EVD prevention behaviours (B = −0.24; 95% CI −0.43, −0.06; P-value = 0.009) [47].

No included publications reported on PTSD and hand hygiene only.

### Combination of disorders and handwashing

Variety existed in the combinations of mental disorders and their associations with handwashing with soap and hand sanitising. When combining anxiety, depression, and PTSD into the single measurement of CMDs, the results showed that while handwashing with soap increased among all participants, the increase level was significantly lower among people with poor mental health than those with good mental health. The influence of mental health on handwashing behaviour was significantly mediated by feelings and difficulty in getting enough soap for handwashing [46].

Four studies combined mental health disorders into a single mental health measure [51-53, 61]. All four studies reported inverse relationships between mental health and handwashing behaviour. During the COVID-19 outbreak in Ethiopia, the odds of having psychological distress was highest among adults who disagree with washing their hands frequently with water and soap to prevent COVID-19 and having the resources to practice such behaviour, (AOR 4.17, 95% CI 1.43-12.15) and (AOR: 2.62; 95% CI 1.20–5.70), respectively [53]. Similar findings were found in Peru, where a statistically significant negative association was found between psychological distress and washing hands after coughing, touching the nose, or sneezing (P-value < 0.001) and washing hands with soap and water (P-value = 0.035) [51]. Similar results were also found in a complex crisis and displacement camp setting.

In rural Malawi, a study found a significant negative association between mental health and handwashing with soap at key times (r = −0.135, P-value ≤ 0.01) after a group of participants received an intervention on the importance of food hygiene techniques. The levels of handwashing with soap at key times in this group was lower among people with poor mental health compared to people with good mental health but the influence of the intervention on handwashing with soap at key times was mediated by mental health (b = 0.0205, CI 0.0035 to 0.0439) [52]. In another non-outbreak setting, participants residing in displacements camp in Northern-Iraq explained that they had been experiencing mental health challenges because of conflict. Some explained that they relied on handwashing as a coping mechanism that helped them manage their trauma and worries, but the associational relationship between mental health challenges and handwashing with soap at critical times varied at an individual level across the different types of camp settings [6 1].

### Examples of Mental Health and Handwashing Programming

Grey literature reports provided case studies from different settings, including anecdotal reports of high distress levels among crisis-affected populations. In a case study of Cox’s Bazar in Bangladesh, psychosocial volunteer investigated available MHPSS for displaced populations. Though many individuals in the camps showed high distress levels, many did not initially prioritize seeking such services because of physical and material needs [55]. Furthermore, the standard of hygiene in such settlements influence people’s health and their psychosocial well-being [45]. Simultaneously, it is often difficult for people suffering from the effects of trauma to devote sufficient attention to personal hygiene. In the case study of Syrian refugees in Jordan, ACF’s mental health and care practices staff identified inadequate hygiene as a contributing factor to psychological distress in refugee populations [45]. These findings were mirrored in a study in IDP camps in Iraqi Kurdistan, where mental health challenges arose as barriers to practicing hand washing [60].

## Discussion

### Summary of findings: analysing the literature

This review aimed to map out what is known about the association between mental health and people’s perceived and actual ability to practice handwashing behaviours in humanitarian crises. The review filled a gap in existing literature as the link between hygiene and psychological health had yet to be extensively studied [20]. Ultimately, 25 publications were included, 21 peer-reviewed articles, and four grey literature publications. Despite the overall lack of a consensus among the findings, some patterns emerged. The most common patterns form the basis for the recommendations for practitioners.

The most common finding for anxiety was that participants with higher rates of anxiety were more likely to practice handwashing with soap. Existing literature reviews and studies of pandemic-related behaviours and psychological outcomes also demonstrated this relationship [50]. Anxious individuals may have higher vigilance levels or concerns about the disease epidemic. It may also be that increased vigilance leads to the development of anxiety in individuals who are more engaged in precautionary measures [47]. This may be because anxiety is elicited primarily by a threat and negative emotions. In general, anxiety may have evolved to serve adaptive and protective functions to help keep individuals safe [63, 64]. In the COVID-19 pandemic, this may be the case where negative emotions and worry encourage adherence to public health-compliant behaviour change [64, 62]. This also supports the findings that decreased anxiety rates led to decreased handwashing rates [56].

Another emerging pattern was contradictory, namely that those with higher rates of anxiety were less likely to wash their hands with soap [48, 49]. This finding was also reported in the existing literature [65, 66]. In Malawi, a complex crisis setting, researchers also found differences between people with higher and lower levels of depression, anxiety, and PTSD in changes to psychosocial factors. These factors included experiencing positive feelings while washing hands with soap [46]. People with poor mental health perceive themselves to be more vulnerable, are less confident about performing preventive behaviours, forget to wash their hands with soap more often, and are less committed to washing their hands with soap [66, 67].

The review found mixed results for the association between handwashing and depression. Four of the seven studies reported that those with higher rates of depression were less likely to wash their hands. In contrast, the remaining studies found that higher depressions scores resulted in more handwashing. Previous research done among children in Zimbabwe supported this inverse relationship between the two factors [18]. Depressed individuals more frequently forgot to wash their hands and experienced less pleasure. They also felt less guilty when not washing their hands and had less intention to wash their hands with soap. Such individuals considered themselves less vulnerable to contracting diseases and were less aware of disease severity. Overall, depression moderates the relationship between behavioural determinants and handwashing through negative patterns of thought, evaluations of the self, the environment, the future, thoughts of worthlessness, and thoughts of death or suicide [18].

PTSD and handwashing also found mixed results. Of the three studies on PTSD, two found that lower scores of PTSD were associated with better hygiene practices, including handwashing with soap. In an outbreak in a post-war setting, higher war exposure was associated with more frequent handwashing. However, PTSD symptoms specifically were associated with less handwashing [47]. Among those having experienced war, individuals with higher exposure to traumatic experiences may have more resilient survival skills or have become more risk averse. Individuals experiencing PTSD may deal with this in the same way, potentially because the salience of the threat of a disease like EVD may not be as great as that of violence and war [47, 68]. Similarly, studies have found that among resettled refugees in high-income countries, the threat of disease may bring up memories of lower standards of hygiene, lack of health infrastructure, and inadequate supplies, which may exacerbate existing mental health conditions [69].

The contradictory patterns suggest that researchers and practitioners need to explore this association further. Improving outcome measurement through standardized and non-self-reported tools and utilizing different study designs utilising qualitative methods could help increase the evidence base. The lack of practical recommendations is also worth exploring and should be the subject of further research and programming. There are opportunities for hygiene programs to strengthen social support mechanisms, for example, by looking to care groups as a potential model. Similarly, hygiene practitioners could be trained in mental health first aid as part of their programming to identify the needs of their target population. Providing simple mental health tips could help individuals deal with their conditions.

### Summary of findings: characteristics of the studies

Most of the research was conducted during the COVID-19 pandemic, likely stemming from the worldwide promotion of handwashing and self-isolation as key preventive measures against infection. Promoting handwashing is typically done in epidemics in the initial absence of vaccines and treatments [57]. The COVID-19 pandemic was unprecedented. A novel coronavirus discovered in Wuhan, China, spread rapidly worldwide, causing widescale mortality, economic devastation, and unprecedented measures, including border closures and restrictions in day-to-day life [62]. The high number of publications from China may be because the virus was first detected in this region. The pandemic has impacted the mental health of the public through the promotion of self-isolation measures which has led to psychological crises in some cases because of rapidly changing rules, fear, and insecurity [70].

Included publications were skewed towards outbreaks, and no included publications reported on natural disasters. Similarly, only a few publications reported on displaced populations as the study population (n = 4). Therefore, it may be difficult to generalize these findings to displaced populations. Despite this, in the publications studying displaced populations, it was clear that dealing with mental health is not always at the forefront of people’s minds [55, 71]. Despite these innovative approaches, a lack of handwashing hardware and challenges accessing the hardware and mental health services still impede handwashing practices in camps [45, 60, 71].

### Methodological limitations of included studies

A series of methodological limitations of included studies exist. The limitations included a lack of qualitative literature in the sample, longitudinal studies and standardized outcome measures for handwashing.

There is a gap in how far an association can be established. Grey literature, as well as some peer-reviewed articles, provided a source of qualitative literature. Here, case studies and interviews provided an insight into the relationship between handwashing and mental health at the individual level.

Similarly, there was a lack of longitudinal studies. Most studies in the analysis were cross-sectional and therefore reflected the psychological state of the study participants at a single period. Psychological states change with time and according to one’s environment [70]. Given the possibility of humanitarian crises to shift into protracted crises, there is a need to investigate the long-term impact of a humanitarian crisis on mental health and handwashing behaviour separately and together.

Lastly, the tools to measure mental health outcome measures and handwashing behaviours were inconsistent across the studies and grey literature publications. This inconsistency creates a limitation in how far an association can be extracted from this data.

### Mediating factors affecting hygiene and handwashing

Upon analysis of the findings, three major themes arose, impacting the links between handwashing and mental health conditions. These include the risk of infection, information exposure, and social support.

#### Risk of infection

The review found that risk of infection was a mediating factor between the association of handwashing and mental health. Many included publications were done in or near the geographical epicentre of the COVID-19 outbreak (n = 12). Residing in the geographical epicentre of an outbreak may have contributed to anxiety mediating the relationship with the practicing of handwashing behaviour due to the increased likelihood of infection. Previous literature [66] supports this finding where living in the geographical epicentre of an outbreak was likely related to an increased perceived risk of infection through family, friends, or community members [70]. The idea is that an increased risk of infection may lead to increased compliance with protective measures, including handwashing, affecting mental health [70]. A similar study found that the top five motivators for handwashing among internally displaced persons were protecting children from illness, ridding the body of germs, avoiding illness, cleanliness, and fear of illness [71]. It is anxiety above normal levels that may make this a self-fulfilling prophecy that weakens the body’s immune system and consequently increases the risk of contracting the virus [70]. Similarly, some participants never felt clean after disinfecting and scrubbing their hands and items repeatedly [42]. These individuals had significantly higher depression scores. Additionally, those who were more concerned about COVID-19 were more engaged in public health-compliance behaviours such as regular handwashing [64].

#### Information exposure

Three publications outlined that COVID-19-related information exposure mediated the association between mental health and handwashing [37, 43, 44]. In one study based in Malawi, mental health mediated the relationship between information exposure and handwashing with soap at key times [52]. Mixed messages from official sources may lead to confusion and fear, allowing anxiety to spread rapidly within families and communities [62, 65, 72]. Emotional contagion is a concept where people transfer negative and positive emotions to others where people who feel threatened, particularly when in crowds, may panic and act to ensure self-preservation [73]. This may explain why people participated more in handwashing behaviour as they are adopting and copying the behaviour of others, acting following public health messaging and social norms [62]. At the same time, however, research has found that following more COVID-19 news leads to more anxiety as such news is distressing, and misinformation can exacerbate depressive symptoms [70]. Media should address low (health) literacy where information should be simple, understandable, and easy to implement in households [68]. Communities, particularly those affected by humanitarian crises or in low-resource settings, should be involved in designing such messaging to adapt such messaging as appropriate [54].

#### Social support

Many publications described social support as a critical protective factor for mental health and associated with a higher frequency of handwashing, mirrored in the existing literature [75]. Social support was conceptualized as providing individuals with stress management training, strengthening coping skills, recommending strategies for maintaining support relationships, and perceived help or care from others [42, 56]. A higher level of social support provided individuals with more cognitive, emotional, and tangible resources to handle the adversities brought on by the epidemic [56]. Existing literature has shown that social support is effective at preventing the development of mental health problems, particularly during a public health crisis [74, 75]. This is because social support and adequate sources of support can help release stress, maintain a person’s emotional responses, and provide positive models for health behaviours by encouraging engagement in personal preventive behaviours [56, 75].

Noncompliance with preventive measures could be a harmful coping response to depressive symptoms, so providing psychological support may be helpful to enhance compliance [77]. Positive emotions and psychological responses such as calmness and optimism are the reasons behind the success of social support as a protective factor. They can reduce such symptoms and, in turn, increase handwashing behaviour [42]. Guidelines for handwashing promotion in the field also mirror this increasing move to positive emotion-based nudging [78].

### Emerging insights for researchers and practitioners

A few critical areas for integration between the WASH and MHPSS sectors are clear. Firstly, humanitarian response should increasingly incorporate mental health [45, 55]. This may be done by incorporating mental health assessments in WASH surveys. The SRQ-20 is an easy screening tool to measure this in the field without requiring psychological training for data collection [46]. Additionally, people with poor mental health should receive treatment before or in parallel with handwashing-promoting interventions to increase positive emotions [46]. Acknowledging that this is extremely difficult to do in a crisis, care may be delivered through adapted technology-based interventions. One publication reported that during the COVID-19 pandemic, WASH and MHPSS teams went door-to-door to distribute hygiene information, soap, and other materials. Addressing mental health during WASH training meant more attention was given to the hygiene session because recipients were less stressed [55]. UNHCR reported similar findings in the field where an elderly displaced Syrian woman experienced traumatic events but with the distribution of a hygiene kit, the stress she experienced compounded the pandemic had subsided [54]. Another intervention could be online or telephone-based Cognitive Behavioural Therapy, which may be used during outbreaks and other humanitarian crises [42, 47, 55, 57] or through the use of digital applications following some specific recommendations [83]. Finally, long-term investments in public, local, and community-based MHPSS should be made as preparedness and resilience help societies better respond to MHPSS needs [55]. Particularly, integration in response plans and MHPSS across sectors should increase access to treatment, ensuring better outcomes across the board [55].

The majority of included studies relied on survey-based quantitative measures to assess mental health and handwashing outcomes. Other, more qualitative methods, such as case studies, ethnographies, focus group discussions, and key informant interviews, may provide a better insight into the perceived barriers to handwashing among those experiencing mental health disorders. Similarly, utilizing more longitudinal approaches could aid in understanding causality in the association and help distinguish chronic mental health disorders from acute disorders.

There were also limitations with the outcomes measures themselves. Studies primarily used self-reported measures of behaviours and used various questions to gauge them. The MHPSS sector should promote a standardized tool to measure each mental health disorder to improve the quality of outcomes measures and standardize this in future research. The most common tools were the GAD-7 for anxiety, the PHQ-9 for Depression, and IES-R for PTSD based on this review. Other literature has also found these tools most common in humanitarian settings [79]. PHQ-9, GAD-7, and IES-R are brief, easy to carry out, and validated in a wide range of settings and populations [79-81]. However, limitations exist, such as overestimating prevalence, lack of cultural sensitivity, and reliance on DSM-IV criteria [79]. Still, these could be promoted as standard tools to measure these disorders in humanitarian settings. This review demonstrates that self-reported questionnaires are most common for handwashing because they are easy to use in low-resource settings. However, to limit social desirability, verification of soap and water availability at dedicated handwashing stations can be used to add a level of validity.

Current approaches to research in humanitarian crises are siloed with mental health and hygiene researchers operating in different spaces. Working groups and communities of practices on WASH and MHPSS can be established to share key learning, best practices from the field, and challenges experienced to bring the two sectors together. Similarly, guiding documents such as ACF’s manual [45] on integrating WASH and Mental Health and Care Practices for humanitarian projects provide concrete ways to integrate the sectors. Sector integration can be carried out joint assessments for WASH and MHPSS interventions, mainstreaming activities by including an additional element in sector-specific programs and integrating with other actors.

Two main gaps were found based on this review. Firstly, the link between mental health and hygiene behaviour in other types of humanitarian crises; other populations, particularly displaced populations; and other countries beyond China should be researched. Handwashing promotion during crises is considered a secondary priority, except in WASH-related disease outbreaks [4]. There is evidence, that when handwashing messages are disseminated, behaviour change initiatives are less effective in acute emergency contexts than relatively stabilized situations or development contexts [78]. This was not explored in the literature and is, therefore, an area of future work. Secondly, there is a need to research specific emotional drivers for practicing handwashing through qualitative approaches, particularly among displaced populations. This is necessary because it will provide an insight into challenges perceived at the individual level.

The dearth of literature overall suggests a greater need for investigation on the link between mental health and people’s perceived and actual ability to practice handwashing. Therefore, a systematic review should not be done. Instead, more field studies are recommended during other types of humanitarian crises and on different populations.

### Limitations of the scoping review

As with all reviews, a few limitations exist. Firstly, the sample size was small and not representative of other humanitarian crises beyond outbreaks, particularly outbreak in China. One reason for this was that only English texts were included. However, the goal of a scoping review is not generalizability but to provide clarity on what is known in a subject area. Furthermore, scoping reviews do not assess the quality of evidence. So, there is a danger that the existence of studies rather than their quality is used as the basis for conclusions. Therefore, these findings cannot be used to recommend policy [82]. The impact of this limitation was reduced through the consultation process with relevant stakeholders and the target of informing relevant WASH and mental health stakeholders on whether a systematic review is needed. Through these processes, relevant experts were able to contribute to this review and determine for themselves the quality of evidence. Moreover, most included studies were cross-sectional, and thus causality was impossible to determine. Other types of literature, such as grey literature, were also consulted to minimize the impact of this, which provided examples of pre-existing and developing mental health disorders and their links to handwashing.

## Conclusion

In conclusion, this review found a lack of a consensus among findings to determine the association between mental health and handwashing. In spite of this, some patterns were more common than others. Particularly among people with anxiety, the likelihood handwashing increased as anxiety levels increased. Among those with depressive- and PTSD symptoms as well as studies measuring a combination of disorders, inverse relationships between mental health disorders and the likelihood of handwashing were more common. Still, there are some emerging insights for practitioners, particularly in integrating mental health assessments in WASH surveys, ensuring that mental health treatment is done in parallel if not before handwashing-promoting interventions, and long-term investments in public, local, and community-based MHPSS to support community resilience. Similarly, research should be continued in this area, notably to explore the link between mental health and people’s perceived and actual ability to practice handwashing, particularly in other types of humanitarian crises; other populations, particularly displaced populations; other countries, beyond China; and at the individual level. The future lies in moving past the siloed nature of humanitarian service provision and only through integrating sectors, can people’s health during humanitarian crises be safeguarded.

## Data Availability

All relevant data are within the manuscript and its Supporting Information files.

## Acknowledgements

We would like to thank Stephanie Stern, and Alan Ricardo Patlán Hernández for providing useful comments on the literature review protocol. We’d like to thank Jennifer Lamb for reviewing the manuscript.

The research was made possible by the generous support of the American people through the United States Agency for international development’s Bureau of Humanitarian Assistance. The contents are the responsibility of the authors of the paper and do not necessarily reflect the views of USAID or the United States Government.

## Supporting Information

**S1 Table: Detailed Search Terms**

## Notes

### Competing Interest Statement

The authors have declared no competing interest.

### Funding Statement

The author(s) received no specific funding for this work. This work was conducted as part of EYG's masters dissertation and was designed to complement the work of the Wash’Em Project. The publication costs were covered by Bureau of Humanitarian Assistance (United States Agency for International Development) who fund Wash’Em (Grant number: 720FDA20GR00351). The funders had no role in study design, data collection and analysis, decision to publish, or preparation of the manuscript.

### Author Declarations

The protocol was reviewed and approved by the ethics committee at the London School of Hygiene and Tropical Medicine (submission ID: 25636).

## References

1. Humanitarian Coalition. What Is a Humanitarian Emergency? [Internet]. 2015 [cited 2021 Apr 9]. Available from: https://www.humanitariancoalition.ca/what-is-a-humanitarian-emergency

2. Shackelford BB, Cronk R, Behnke N, Cooper B, Tu R, D’Souza M, et al. Environmental health in forced displacement: A systematic scoping review of the emergency phase. Sci Total Environ. 2020 Apr 20;714:136553.

3. Phillips RM, Vujcic J, Boscoe A, Handzel T, Aninyasi M, Cookson ST, et al. Soap is not enough: handwashing practices and knowledge in refugee camps, Maban County, South Sudan. Conflict and Health. 2015 Dec 20;9(1):39.

4. Vujcic J, Ram PK, Blum LS. Handwashing promotion in humanitarian emergencies: strategies and challenges according to experts. Journal of Water, Sanitation and Hygiene for Development. 2015 Sep 23;5(4):574–85.

5. Checchi F, Warsame A, Treacy-Wong V, Polonsky J, Ommeren M van, Prudhon C. Public health information in crisis-affected populations: a review of methods and their use for advocacy and action. The Lancet. 2017 Nov 18;390(10109):2297–313.

6. Connolly MA, Gayer M, Ryan MJ, Salama P, Spiegel P, Heymann DL. Communicable diseases in complex emergencies: impact and challenges. Lancet. 2004 Dec 27;364(9449):1974–83.

7. Médecins Sans Frontières. Treating diarrhoea in emergency settings | MSF [Internet]. Médecins Sans Frontières (MSF) International. 2003 [cited 2021 Aug 28]. Available from: https://www.msf.org/treating-diarrhoea-emergency-settings

8. Aiello AE, Larson EL. What is the evidence for a causal link between hygiene and infections? Lancet Infect Dis. 2002 Feb;2(2):103–10.

9. Rabie T, Curtis V. Handwashing and risk of respiratory infections: a quantitative systematic review. Trop Med Int Health. 2006 Mar;11(3):258–67.

10. Wolf J, Hunter PR, Freeman MC, Cumming O, Clasen T, Bartram J, et al. Impact of drinking water, sanitation and handwashing with soap on childhood diarrhoeal disease: updated meta-analysis and meta-regression. Tropical Medicine & International Health. 2018;23(5):508–25.

11. Freeman MC, Stocks ME, Cumming O, Jeandron A, Higgins JPT, Wolf J, et al. Hygiene and health: systematic review of handwashing practices worldwide and update of health effects. Trop Med Int Health. 2014 Aug;19(8):906–16.

12. Ejemot-Nwadiaro RI, Ehiri JE, Arikpo D, Meremikwu MM, Critchley JA. Hand washing promotion for preventing diarrhoea. Cochrane Database Syst Rev. 2015 Sep 3;(9):CD004265.

13. Curtis V, Cairncross S. Effect of washing hands with soap on diarrhoea risk in the community: a systematic review. Lancet Infect Dis. 2003 May;3(5):275–81.

14. White S, Thorseth AH, Dreibelbis R, Curtis V. The determinants of handwashing behaviour in domestic settings: An integrative systematic review. International Journal of Hygiene and Environmental Health. 2020 Jun 1;227:113512.

15. World Health Organization. Mental health: strengthening our response [Internet]. World Health Organization. 2018 [cited 2021 Mar 15]. Available from: https://www.who.int/news-room/fact-sheets/detail/mental-health-strengthening-our-response

16. Silove D, Ventevogel P, Rees S. The contemporary refugee crisis: an overview of mental health challenges. World Psychiatry. 2017 Jun;16(2):130–9.

17. Charlson F, Ommeren M van, Flaxman A, Cornett J, Whiteford H, Saxena S. New WHO prevalence estimates of mental disorders in conflict settings: a systematic review and meta-analysis. The Lancet. 2019 Jul 20;394(10194):240–8.

18. Slekiene J, Mosler HJ. Does depression moderate handwashing in children? BMC Public Health. 2018;18(1):82.

19. Fontenelle LF, Cocchi L, Harrison BJ, Shavitt RG, do Rosário MC, Ferrão YA, et al. Towards a post-traumatic subtype of obsessive–compulsive disorder. Journal of Anxiety Disorders. 2012 Mar 1;26(2):377–83.

20. Ranasinghe S, Ramesh S, Jacobsen KH. Hygiene and mental health among middle school students in India and 11 other countries. Journal of Infection and Public Health. 2016 Jul 1;9(4):429–35.

21. Stewart V, Judd C, Wheeler AJ. Practitioners’ experiences of deteriorating personal hygiene standards in people living with depression in Australia: A qualitative study. Health Soc Care Community. 2021 Jul 8;

22. Stevenson RJ, Case TI, Hodgson D, Porzig-Drummond R, Barouei J, Oaten MJ. A scale for measuring hygiene behavior: development, reliability and validity. Am J Infect Control. 2009 Sep;37(7):557–64.

23. Leibold ML, Holm MB, Raina KD, Reynolds CF, Rogers JC. Activities and adaptation in late-life depression: a qualitative study. Am J Occup Ther. 2014 Oct;68(5):570–7.

24. Contzen N, De Pasquale S, Mosler HJ. Over-Reporting in Handwashing Self-Reports: Potential Explanatory Factors and Alternative Measurements. PLoS One. 2015 Aug 24;10(8):e0136445.

25. Ram P. Practical Guidance for Measuring Handwashing Behavior. Water and Sanitation Program : working paper [Internet]. Washington DC: World Bank; 2010 p. 16. Available from: http://hdl.handle.net/10986/19005

26. Halder AK, Molyneaux JW, Luby SP, Ram PK. Impact of duration of structured observations on measurement of handwashing behavior at critical times. BMC Public Health. 2013 Aug 2;13(1):705.

27. Arksey H, O’Malley L. Scoping studies: towards a methodological framework. International Journal of Social Research Methodology. 2005 Feb 1;8(1):19–32.

28. American Psychiatric Association. Diagnostic and statistical manual of mental disorders [Internet]. 5th ed. Washington DC; 2013 [cited 2021 Jun 23]. Available from: https://www.ptsd.va.gov/professional/treat/essentials/dsm5_ptsd.asp#one

29. Paludan-Müller AS, Boesen K, Klerings I, Jørgensen KJ, Munkholm K. Hand cleaning with ash for reducing the spread of viral and bacterial infections: a rapid review. Cochrane Database of Systematic Reviews [Internet]. 2020 [cited 2021 Aug 21];(4). Available from: https://www.cochranelibrary.com/cdsr/doi/10.1002/14651858.CD013597/full

30. United Nations High Commissioner for Refugees. Refugee Data Finder [Internet]. UNHCR. 2021 [cited 2021 Mar 18]. Available from: https://www.unhcr.org/refugee-statistics/

31. Hamadeh N, Van Rompaey C, Metreau E. New World Bank country classifications by income level: 2021-2022 [Internet]. World Bank Blogs. 2021 [cited 2021 Jul 15]. Available from: https://blogs.worldbank.org/opendata/new-world-bank-country-classifications-income-level-2021-2022

32. Hoque BA. Handwashing practices and challenges in Bangladesh. Int J Environ Health Res. 2003 Jun;13 Suppl 1:S81–87.

33. Prajapati P, Desai H, Chandarana C. Hand sanitizers as a preventive measure in COVID-19 pandemic, its characteristics, and harmful effects: a review. Journal of the Egyptian Public Health Association. 2022 Feb 8;97(1):6.

34. World Health Organization. Advice for the public on COVID-19 – World Health Organization [Internet]. Advice for the public: Coronavirus disease (COVID-19). 2023 [cited 2023 Apr 16]. Available from: https://www.who.int/emergencies/diseases/novel-coronavirus-2019/advice-for-public

35. Jess RL, Dozier CL. Increasing handwashing in young children: A brief review. Journal of Applied Behavior Analysis. 2020;53(3):1219–24.

36. Substance Abuse and Mental Health Services Administration. Impact of the DSM-IV to DSM-5 Changes on the National Survey on Drug Use and Health [Internet]. Rockville (MD): Substance Abuse and Mental Health Services Administration (US); 2016 [cited 2021 Jun 23]. (CBHSQ Methodology Report). Available from: http://www.ncbi.nlm.nih.gov/books/NBK519697/

37. Qian M, Wu Q, Wu P, Hou Z, Liang Y, Cowling BJ, et al. Anxiety levels, precautionary behaviours and public perceptions during the early phase of the COVID-19 outbreak in China: a population-based cross-sectional survey. BMJ Open. 2020 Oct 1;10(10):e040910.

38. Goodwin R, Sun S. Early responses to H7N9 in southern Mainland China. BMC Infect Dis. 2014 Jan 7;14:8.

39. Mohammadpour M, Ghorbani V, Khoramnia S, Ahmadi SM, Ghvami M, Maleki M. Anxiety, Self-Compassion, Gender Differences and COVID-19: Predicting Self-Care Behaviors and Fear of COVID-19 Based on Anxiety and Self-Compassion with an Emphasis on Gender Differences. Iran J Psychiatry. 2020 Jul;15(3):213–9.

40. Wang J, Rao N, Han B. Pathways Improving Compliance with Preventive Behaviors during the Remission Period of the COVID-19 Pandemic. International Journal of Environmental Research and Public Health. 2021 Jan;18(7):3512.

41. Birhanu A, Tiki T, Mekuria M, Yilma D, Melese G, Seifu B. COVID-19-Induced Anxiety and Associated Factors Among Urban Residents in West Shewa Zone, Central Ethiopia, 2020. Psychol Res Behav Manag. 2021 Feb 9;14:99–108.

42. Zhang W, Yang X, Zhao J, Yang F, Jia Y, Cui C, et al. Depression and Psychological-Behavioral Responses Among the General Public in China During the Early Stages of the COVID-19 Pandemic: Survey Study. J Med Internet Res. 2020 Sep 4;22(9):e22227.

43. Pan Y, Fang Y, Xin M, Dong W, Zhou L, Hou Q, et al. Self-Reported Compliance With Personal Preventive Measures Among Chinese Factory Workers at the Beginning of Work Resumption Following the COVID-19 Outbreak: Cross-Sectional Survey Study. J Med Internet Res. 2020 Sep 29;22(9):e22457.

44. Pan Y, Xin M, Zhang C, Dong W, Fang Y, Wu W, et al. Associations of Mental Health and Personal Preventive Measure Compliance With Exposure to COVID-19 Information During Work Resumption Following the COVID-19 Outbreak in China: Cross-Sectional Survey Study. Journal of Medical Internet Research. 2020 Oct 8;22(10):e22596.

45. Action contre la Faim. 1 + 1 = 3: How to Integrate WASH and MHCP Activities for Better Humanitarian Projects [Internet]. France: Action contre la Faim; 2013 Dec [cited 2021 Aug 27] p. 74. Available from: https://www.actioncontrelafaim.org/publication/acf-international-manual-1-1-3-how-to-integrate-wash-and-mhcp-activities-for-better-humanitarian-projects/

46. Slekiene J, Mosler HJ. Does poor mental health change the influence of interventions on handwashing in a vulnerable population of rural Malawi? The key role of emotions. Journal of Water, Sanitation and Hygiene for Development. 2020 Dec 30;11(3):350– 61.

47. Betancourt TS, Brennan RT, Vinck P, VanderWeele TJ, Spencer-Walters D, Jeong J, et al. Associations between Mental Health and Ebola-Related Health Behaviors: A Regionally Representative Cross-sectional Survey in Post-conflict Sierra Leone. PLoS Med. 2016 Aug;13(8):e1002073.

48. Wang C, Pan R, Wan X, Tan Y, Xu L, McIntyre RS, et al. A longitudinal study on the mental health of general population during the COVID-19 epidemic in China. Brain, Behavior, and Immunity. 2020 Jul 1;87:40–8.

49. Wang C, Pan R, Wan X, Tan Y, Xu L, Ho CS, et al. Immediate Psychological Responses and Associated Factors during the Initial Stage of the 2019 Coronavirus Disease (COVID-19) Epidemic among the General Population in China. Int J Environ Res Public Health. 2020 Mar 6;17(5):E1729.

50. Assefa ZM, Haile TG, Wazema DH, Tafese WT, Berrie FW, Beketie ED, et al. Mental Health Disorders During COVID-19 Pandemic Among Southwest Ethiopia University Students: An Institutional-Based Cross-Sectional Study. SAGE Open Nurs. 2021;7:23779608211064376.

51. Ruiz-Frutos C, Palomino-Baldeón JC, Ortega-Moreno M, Villavicencio-Guardia M del C, Dias A, Bernardes JM, et al. Effects of the COVID-19 Pandemic on Mental Health in Peru: Psychological Distress. Healthcare. 2021 Jun;9(6):691.

52. Slekiene J, Chidziwisano K, Morse T. Does Poor Mental Health Impair the Effectiveness of Complementary Food Hygiene Behavior Change Intervention in Rural Malawi? International Journal of Environmental Research and Public Health. 2022 Jan;19(17):10589.

53. Ambelu A, Birhanu Z, Yitayih Y, Kebede Y, Mecha M, Abafita J, et al. Psychological distress during the COVID-19 pandemic in Ethiopia: an online cross-sectional study to identify the need for equal attention of intervention. Annals of General Psychiatry. 2021 Mar 25;20(1):22.

54. United Nations High Commissioner for Refugees. Emerging Practices: Mental health and psychosocial support in refugee operations during the COVID-19 pandemic - World [Internet]. Geneva: United Nations High Commissioner for Refugees; 2020 Jun [cited 2021 Aug 27] p. 16. Available from: https://reliefweb.int/report/world/emerging-practices-mental-health-and-psychosocial-support-refugee-operations-during

55. Clomén D, Horn R, Osorio M, Powell M, Rydell AF, Sejberg Z. “The greatest need was to be listened to”: The importance of mental health and psychosocial support during COVID-19 [Internet]. Geneva: International Committee of the Red Cross and International Federation of Red Cross and Red Crescent Societies; 2020 Oct [cited 2021 Aug 27] p. 23. Available from: https://pscentre.org/?resource=the-greatest-need-was-to-be-listened-to-the-importance-of-mental-health-and-psychosocial-support-during-covid-19&selected=single-resource

56. Wang Q, Mo PKH, Song B, Di JL, Zhou FR, Zhao J, et al. Mental health and preventive behaviour of pregnant women in China during the early phase of the COVID-19 period. Infectious Diseases of Poverty. 2021 Mar 24;10(1):37.

57. Ni Z, Lebowitz ER, Zou Z, Wang H, Liu H, Shrestha R, et al. Response to the COVID-19 Outbreak in Urban Settings in China. J Urban Health. 2021 Feb;98(1):41–52.

58. Liu X, Luo WT, Li Y, Li CN, Hong ZS, Chen HL, et al. Psychological status and behavior changes of the public during the COVID-19 epidemic in China. Infectious Diseases of Poverty. 2020 May 29;9(1):58.

59. Liu J, Tong Y, Li S, Tian Z, He L, Zheng J. Compliance with COVID-19-preventive behaviours among employees returning to work in the post-epidemic period. BMC Public Health. 2022 Feb 21;22(1):369.

60. Zangana A, Shabila N, Heath T, White S. The determinants of handwashing behaviour among internally displaced women in two camps in the Kurdistan Region of Iraq. PLOS ONE. 2020 May 8;15(5):e0231694.

61. White S, Heath T, Ibrahim WK, Ihsan D, Blanchet K, Curtis V, et al. How is hygiene behaviour affected by conflict and displacement? A qualitative case study in Northern Iraq. PLOS ONE. 2022 Mar 3;17(3):e0264434.

62. Usher K, Jackson D, Durkin J, Gyamfi N, Bhullar N. Pandemic-related behaviours and psychological outcomes; A rapid literature review to explain COVID-19 behaviours. International Journal of Mental Health Nursing. 2020;29(6):1018–34.

63. Perkins AM, Corr PJ. Anxiety as an adaptive emotion. In: The positive side of negative emotions. New York, NY, US: The Guilford Press; 2014. p. 37–54.

64. Harper CA, Satchell LP, Fido D, Latzman RD. Functional Fear Predicts Public Health Compliance in the COVID-19 Pandemic. Int J Ment Health Addict. 2020 Apr 27;1–14.

65. Lin CY, Lin YL. Anxiety and depression of general population in the early phase of COVID-19 pandemic: A systematic review of cross-sectional studies. Arch Clin Psychiatry (São Paulo). 2021;47:199–208.

66. Slekiene J, Mosler HJ. Poor mental health impairs WASH behaviors: Including people with poor mental health in WASH interventions. [Internet]. Dübendorf, Switzerland: Eawag, Swiss Federal Institute of 2 Aquatic Science and Technology; 2019 Oct p. 2. (Intervention Fact Sheet 9: Data-Driven Behavior Change). Available from: https://www.ranasmosler.com/mental-health

67. Schiavo R, May Leung M, Brown M. Communicating risk and promoting disease mitigation measures in epidemics and emerging disease settings. Pathog Glob Health. 2014 Mar;108(2):76–94.

68. Coetzee BJ, Kagee A. Structural barriers to adhering to health behaviours in the context of the COVID-19 crisis: Considerations for low- and middle-income countries. Global Public Health. 2020 Aug 2;15(8):1093–102.

69. Brickhill-Atkinson M, Hauck FR. Impact of COVID-19 on Resettled Refugees. Prim Care. 2021 Mar;48(1):57–66.

70. Salari N, Hosseinian-Far A, Jalali R, Vaisi-Raygani A, Rasoulpoor S, Mohammadi M, et al. Prevalence of stress, anxiety, depression among the general population during the COVID-19 pandemic: a systematic review and meta-analysis. Globalization and Health. 2020 Jul 6;16(1):57.

71. Blum LS, Yemweni A, Trinies V, Kambere M, Tolani F, Allen JV, et al. Programmatic implications for promotion of handwashing behavior in an internally displaced persons camp in North Kivu, Democratic Republic of Congo. Conflict and Health. 2019 Nov 20;13(1):54.

72. Kramer ADI, Guillory JE, Hancock JT. Experimental evidence of massive-scale emotional contagion through social networks. PNAS. 2014 Jun 17;111(24):8788–90.

73. Clarke L. Panic: Myth or Reality? Contexts. 2002 Aug 1;1(3):21–6.

74. Wang X, Cai L, Qian J, Peng J. Social support moderates stress effects on depression. Int J Ment Health Syst. 2014 Jan 1;8(1):41.

75. Fung ICH, Cairncross S. How often do you wash your hands? A review of studies of hand-washing practices in the community during and after the SARS outbreak in 2003. Int J Environ Health Res. 2007 Jun;17(3):161–83.

76. Maulik PK, Eaton WW, Bradshaw CP. The Role of Social Network and Support in Mental Health Service Use: Findings From the Baltimore ECA Study. PS. 2009 Sep 1;60(9):1222–9.

77. Folkman S, Chesney MA, Pollack L, Phillips C. Stress, coping, and high-risk sexual behavior. Health Psychology. 1992;11(4):218–22.

78. Ramos M, Benelli P, Irvine E, Watson J. WASH in Emergencies Problem Exploration Report: Handwashing [Internet]. Elhra and Humanitarian Innovation Fund; 2016 Jan [cited 2021 Aug 28]. Available from: https://www.elrha.org/researchdatabase/handwashing/

79. Moore A, van Loenhout JAF, de Almeida MM, Smith P, Guha-Sapir D. Measuring mental health burden in humanitarian settings: a critical review of assessment tools. Glob Health Action. 13(1):1783957.

80. Spitzer RL, Kroenke K, Williams JBW, Löwe B. A Brief Measure for Assessing Generalized Anxiety Disorder: The GAD-7. Archives of Internal Medicine. 2006 May 22;166(10):1092–7.

81. Bian C, Li C, Duan Q, Wu H. Reliability and validity of patient health questionnaire: Depressive syndrome module for outpatients. SRE. 2011 Jan 18;6(2):278–82.

82. Grant M, Booth A. A typology of reviews: An analysis of 14 review types and associated methodologies. Health information and libraries journal. 2009 Jul 1;26:91– 108.

83. Ratheesh, A., Alvarez-Jimenez, M. The future of digital mental health in the post-pandemic world: Evidence-based, blended, responsive and implementable. Australian & New Zealand Journal of Psychiatry 2022;56(2):107–109. doi:10.1177/00048674211070984

